# Characteristics of admissions for Kawasaki Disease from 1997 to 2012: Lessons from a national database

**DOI:** 10.1101/2022.04.30.22274523

**Authors:** Alexander Raskin, Rohit S. Loomba

## Abstract

A majority of large epidemiologic studies on Kawasaki Disease have come from Asia. There is paucity of data assessing Kawasaki Disease on a national level in the U.S., particularly in terms of hospitalization co-morbidities and cost. This study set forth to analyze data from the Kids’ Inpatient Database from 1997 to 2012. Data were analyzed for age, race, cardiogenic shock, acute kidney injury, liver failure, acute respiratory distress syndrome, arrhythmia, and congenital heart disease. Additionally, multivariate regression analysis was performed to assess the impact of Kawasaki Disease on coronary artery aneurysms, ECMO, length of stay, cost of stay, and mortality. Asian and Pacific Islander children were disproportionally affected by Kawasaki Disease in the U.S (20.8% of Kawasaki Disease admissions vs 3.3% of all other pediatric hospital admissions, p<0.01). Patients hospitalized for Kawasaki Disease had an increased risk of developing coronary artery aneurysms (OR 2,839, 95%CI 2,2985-3,527) and cardiogenic shock (OR 3.42, 95% CI 2.18-5.37). Patients with Kawasaki disease were less likely to have congenital heart disease (OR 0.62, 95%CI 0.55-0.69), arrhythmia (OR 0.31, 95%CI 0.11-0.84), and acute respiratory distress syndrome (OR 0.29, 95%CI 0.19-0.43). Kawasaki disease patients had shorter hospitalization length of stay by 2.59 days (p <0.01) and decreased cost of stay by $5,513 (p <0.01). Kawasaki Disease had lower mortality when compared to all other admissions (OR 0.03, 95%CI 0.01-0.09). No significant differences were found for ECMO, acute kidney injury, or liver failure.

## Introduction

Kawasaki disease (KD) is the leading cause of acquired pediatric heart disease.^[1,2]^ The diagnosis of KD is based on clinical criteria including: fever, bilateral nonexudative conjunctivitis, erythema of the oral mucosa and lips, rash, cervical lymphadenopathy, and changes in the extremities. ^[3]^ The disease process is usually self-limited, but failure of timely diagnosis and treatment may lead to long term sequelae of which coronary artery aneurysms are of greatest concern. In those who are untreated, coronary artery aneurysms or coronary ectasia develop in 15 to 25% of children. ^[4,5]^ The mainstay of treatment is IVIG and aspirin, which have been shown to reduce the incidence of coronary artery aneurysms. ^[6]^ Unfortunately, not all children respond to IVIG, which poses a management dilemma. ^[7-10]^ Multiple large scale epidemiological studies have been geared towards better understanding the etiology and improving therapeutic interventions. The majority of these studies hail from Japan, South Korea, and Taiwan, where the incidence rates of KD are significantly higher than in the United States.^[11-15]^ Studies assessing KD on a national level in the United States are lacking. This is of significance as previous work has suggested ethnicity based variability in KD.^[13,16,17]^ The aim of this study was to better characterize KD hospitalizations in the United States using nationally available reported data.

## Methods

We analyzed hospital administrative data from the Kids’ Inpatient Database (KID). The KID is a national all payer hospital pediatric database that utilizes discharge abstracts created by hospitals for billing. It is the largest publicly available pediatric inpatient database in the United States. Data from the 1997, 2000, 2003, 2006, 2009, and 2012 were analyzed for the purposes of this study.

Hospitalizations with the diagnosis of KD were identified using International Classification of Diseases, Ninth Revisions, Clinical Modification (ICD-9CM) code 446.1 (mucocutaneous lymph node disease). Patient characteristics were assessed for age and race (Caucasian, African American, Hispanic, Asian/Pacific Islander, or Other).

Admissions with and wthout KD were compared. Univariate analyses were done using student T-tests or Mann-Whitney-U tests for continuous variables and chi-squared tests for descriptive variables based on the normalcy of the data. Regression analyses were then conducted to model the impact of various patient characteristics and comorbidities on hospital characteristics such as duration and cost of stay. These regressions were conducted with the hospital characteristic of interest as the dependent variable and then patient characteristics and comorbidities, including KD, serving as the independent variables.

All statistical analyses were done using SPSS Version 21.0 (Chicago, IL). A p-value of less than 0.05 was considered statistically significant.

## Results

A total of 12,931,595 patients were identified and included for analyses. Of the whole group, 17,734 patients had an ICD-9CM diagnosis code for Kawasaki disease. The remaining 12, 913,861 admissions had a diagnosis other that Kawasaki Disease. The average age of admission for the KD group was 2 years vs. 0.1 years for the non KD group, which was statistically significant (p < 0.01). The race characteristics between the two groups were also statistically significant with a p < 0.01. For the KD group 44.7% were Caucasian, 20.8% were Asian/Pacific Islander, 20.8% were Hispanic, 18.4% were African American, 0.6% were Native American, and 6.2% were identified as Other. For the non Kawasaki group, Caucasians made up 50.9% of the group. Only 3.3% were Asian/Pacific Islander. African American, Hispanic, Native American, and Other made up 17.1%, 22.5%, 0.7%, and 5.5%, respectively, the remainder of the non Kawasaki cohort.

Univariate analysis demonstrated that patients with KD had increased risk of cardiogenic shock, odds ratio of 3.42 (95% CI 2.18-5.37, p <0.01). There was no statistically significant difference for AKI and liver failure between the two groups, odds ratios of 1.25 (95%CI 0.98-1.59) and 1.29 (95%CI 0.71-2.33), respectively. There was statistically significantly less ARDS in the Kawasaki group, odds ratio 0.29 (95%CI 0.19-0.43, p <0.01). There was also significantly less arrhythmia in the Kawasaki group, odds ratio 0.31 (95%CI 0.11-0.84, p = 0.01). Patients with KD were less likely to have congenital heart disease, odds ratio 0.62 (95%CI 0.55-0.69, p <0.01).

Multivariate regression analysis demonstrated that KD increases coronary artery aneurysm risk by an odds of 2,839 (95%CI 2,2985-3,527), p <0.01. Additionally by multivariate regression analysis, Kawasaki disease has shorter hospitalization length of stay by 2.59 days (p <0.01) and decreased cost of stay by $5,513 (p <0.01). KD has lower mortality when compared to all other admissions, odds ratio 0.03 (95%CI 0.01-0.09), p <0.01. There was no statistically significant difference for ECMO between the two groups.

## Discussion

Kawasaki Disease affects children across the globe. However, the epidemiology is variable around the world. As previously described, the highest prevalence of the disease is in Asia. Japan, South Korea, and Taiwan have well established KD registries. From these registries, the annual incidence rates were up to 264.8 per 100,000 in Japan, 113.1 per 100,000 in South Korea, and up to 68.5 per 100,000 in Taiwan.^[11,13,14,18]^ Since 1976, passive surveillance has been conducted on KD in the United States. This data has been supplemented by periodic hospital discharge records. Based on that data, the annual incidence rate of KD in the United States is approximately 19 per 100,000, with higher incidence rates reported amongst Asian and Pacific Islander children.^[14-17]^ This study compiled data from all available KID registries (1997, 2000, 2003, 2006, 2009, and 2012), making it the largest collection of data on KD in the United States. Our findings confirm that Asian and Pacific Islander children are disproportionally affected by KD in the United States, making up 20.8% of all KD admissions in the United States while accounting for only 3.3% of the remaining pediatric hospital admissions. Of greatest concern is if certain ethnicities convey an increased risk for developing coronary artery changes. Our data indicates that 2.7% of children hospitalized for KD developed coronary artery aneurysms, which is an increased risk by an odds of 2,839. This is consistent with previous research that demonstrated coronary artery aneurysm rates between 2.25-4% in the United States KD population.^[13,15]^ Using KID registry data from 2003, 2006, 2009, and 2012, Okubo et al. demonstrated that there were no racial differences in the development of coronary artery aneurysms.^[15]^ However, previous literature has demonstrated that African Americans have higher rates of IVIG resistance.^[19-21]^ Patients that have IVIG resistance, defined as persistence of fevers after the first dose of IVIG, have been shown to have an increased risk of developing coronary artery abnormalities.^[22,23]^ Further research is needed to determine if racial differences in IVIG resistance have an implication on the development of coronary artery aneurysms. To date there is no reliable clinical tool that risk stratifies IVIG resistance or development of coronary artery aneurysms in the U.S KD population. Whereas, Japanese studies have been able to predict IVIG resistance with fairly high sensitivity and specificity.^[24-26]^ Those predicted to be non responders were shown to have reduced rates of coronary artery lesions if they were initially treated with IVIG, aspirin, and pulse steroids vs. those not receiving steroids.^[27]^ Unfortunately, when applied to the U.S. population, these Japanese scoring systems failed to be validated.^[21,28,29]^ National data registries should continue to be utilized in order to help identify improved therapeutic interventions and risk factors associated with the development of coronary artery aneurysms.

As part of this study we were also able to assess other KD co-morbidities at the time of admission. Children with KD had a decreased risk of developing ARDS or an arrhythmia during the time of their admission. However, there was an increased risk of KD patients developing cariogenic shock. The presumed mechanism for this is that KD leads to diffuse myocardial inflammation.^[3]^ Interestingly enough, our data demonstrates that despite conveying a higher risk for cariogenic shock, patients with KD did not have increased ECMO utilization. However, this may be a reflection of limited numbers as only 19 out of the 17,734 children with KD developed cardiogenic (0.1%), and less than 10 children with KD required ECMO support. It is also interesting to note that our data demonstrates that 311 patients (1.8%) with KD had an underlying congenital heart defect (CHD). For unclear reasons, children with CHD had a decreased risk of developing KD. It maybe that certain syndromes and genetic conditions associated with specific CHDs may alter immune response or immune modulation and thus decrease the risk of developing KD. With this current dataset it is unknown if specific CHDs were over or under represented in the KD group. Additionally, literature on the incidence of KD in the CHD population is lacking.

Another aspect to consider when analyzing KD admissions is cost and resource utilization. This is of particular importance in the modern era of medicine. Working with administrative databases gave us a unique opportunity to perform such an analysis. KD admissions had shorter length of stay, lower cost of hospitalization, and lower mortality when compared to all other causes of pediatric hospital admissions. Future studies should tease out whether IVIG resistance significantly affects length of stay and cost. Additionally, this analysis does not take into account mortality, morbidity, and cost after hospital discharge. Frequency and duration of long term follow up is determined by risk level based on degree of coronary artery involvement. Patients with residual coronary artery changes require long term thromboprophylaxis. For patients with residual coronary artery lesions there is increased risk of long term sequelae, such as coronary artery thrombosis. Whether KD patients who have normal coronaries have an increased risk for endovascular disease later in life remains a controversial and debated topic.^[3,30]^

There are several limitations to this study. This was a retrospective study based on administrative data. It is possible that there could have been miscoding or misclassification that could have led to either over or underdiagnoses. Additionally, there were no laboratory, imaging, or detailed clinical information provided to further assess differences between groups. However, the advantage of using these data sets is that it allows for an assessment of KD on a national level with a large sample size.

**Table 1.** Comparison of admissions with and without KD

|  | No KD<br>(n=12,913,861) | KD<br>(n=17,734) | Odds ratio (95% confidence interval) | p-value |
| --- | --- | --- | --- | --- |
| **Age (years)** | 0.1 (0 to 17) | 2.0 (0 to 17) | -- | < 0.01 |
| **Race** |  |  | -- | < 0.01 |
| *Caucasian* | 5,303,386 (50.9) | 6,347 (44.7) |  |  |
| *African American* | 1,782,431 (17.1) | 2,612 (18.4) |  |  |
| *Hispanic* | 2,348,384 (22.5) | 2,959 (20.8) |  |  |
| *Asian or Pacific Islander* | 344,031 (3.3) | 1,330 (20.8) |  |  |
| *Native American* | 77,957 (0.7) | 79 (0.6) |  |  |
| *Other* | 570,220 (5.5) | 882 (6.2) |  |  |
| **Coronary artery aneurysm** | 132 (< 0.1) | 480 (2.7) | 2,722.36 (2,244.08 to 3,302.58) | < 0.01 |
| **Cardiogenic shock** | 4,047 (< 0.1) | 19 (0.1) | 3.42 (2.18 to 5.37) | < 0.01 |
| **Acute kidney injury** | 38,943 (0.3) | 67 (0.4) | 1.25 (0.98 to 1.59) | 0.06 |
| **Liver failure** | 6,198 (< 0.1) | 11 (0.1) | 1.29 (0.71 to 2.33) | 0.39 |
| **Acute respiratory distress syndrome** | 59,838 (0.5) | 24 (0.1) | 0.29 (0.19 to 0.43) | < 0.01 |
| **Arrhythmia (not including atrioventricular block)** | 9,224 (0.1) | *** (***) | 0.31 (0.11 to 0.84) | 0.01 |
| **Congenital heart disease** | 361,230 (2.8) | 311 (1.8) | 0.62 (0.55 to 0.69) | < 0.01 |
| **Length of hospital stay (days)** | 2 (0 to 460) | 3 (0 to 76) | -- | < 0.01 |
| **Cost of hospitalization (US dollars)** | 6,471 | 15,150 | -- | 0.08 |
| **Extracorporeal membrane oxygenation** | 5,783 (0.1) | *** (***) | 0.71 (0.29 to 1.71) | 0.44 |
| **Inpatient mortality** | 88,066 (0.7) | *** (***) | 0.03 (0.01 to 0.08) | < 0.01 |

KD- Kawasaki disease
\*\*\* Represents an absolute frequency less than 10 which per database policies cannot be explicitly reported

**Table 2.** Impact of KD on specific outcomes

| Impact of KD on specific outcomes |  |  |
| --- | --- | --- |
|  | Beta Coefficient or odds ratio (95%CI) | p-value |
| **Coronary artery aneurysm** | 2,839.71 (2,285.77 to 3,527.89) | < 0.01 |
| **ECMO** | 0.95 (0.38 to 2.36) | 0.92 |
| **Length of stay** | -2.59 | < 0.01 |
| **Cost of stay** | -5,513 | < 0.01 |
| **Mortality** | 0.03 (0.01 to 0.09) | < 0.01 |

## Data Availability

All data produced are present

